# Light propagation within N95 Filtered Face Respirators: A simulation study for UVC decontamination

**DOI:** 10.1101/2020.06.12.20129395

**Authors:** Lothar Lilge, Angelica Manalac, Madrigal Weersink, Fynn Schwiegelshohn, Tanner Young-Schultz, Abdallatif Satti Abdalrhman, Chengjin Wang, Aldrich Ngan, Frank X. Gu, Vaughn Betz, Ron Hofmann

## Abstract

This study presents numerical simulations of UVC light propagation through seven different filtered face respirators (FFR) to determine their suitability for UV germicidal inactivation (UVGI). UV propagation was modelled using the FullMonte program for two external light illuminations. The optical properties of the dominant three layers were determined using the inverse adding doubling method.

The resulting fluence rate volume histograms and the lowest fluence rate recorded in the modelled volume, sometimes in the nW cm^-2^, provide feedback on a respirator’s suitability for UVGI and the required exposure time for a given light source. While UVGI can present an economical approach to extend an FFR’s useable lifetime, it requires careful optimization of the illumination setup and selection of appropriate respirators.

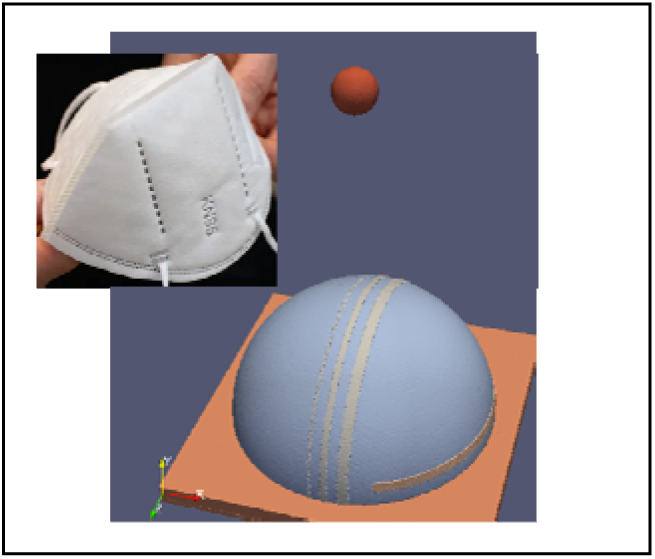

## 1 |INTRODUCTION

Rapid local or global outbreaks of disease, such as SARS, Ebola and COVID-19 caused by SARS-CoV-2, cannot be forecast despite the general knowledge that they will sporadically occur. Health care providers are particularly at risk during such outbreaks. For example, out of the 8096 reported cases during the 2002-2004 SARS outbreak, 1706 (21%) were health care workers [1], as were 221 of the 3956 fatalities in Sierra Leone during the 2019-2020 Ebola outbreak [2]. The death rate associated with SARS-CoV-2 induced COVID-19 ranges from 1.3% (Russia) to 15.5% (France) June 8^th^, 2020 [3] and an average mortality rate of 3.7%, underscoring the need for a very high level of protection for health care workers. Hence, the use of N95 disposable filtering facepiece respirators (FFR) is restricted to front line health care professionals in most North American jurisdictions. Regular face masks are recommended for the broader public to reduce the infection rate further.

The current COVID-19 pandemic has severely stressed supplies of personal protective equipment (PPE), including N95 filtering facepiece respirators (N95 FFRs). In response, healthcare facilities worldwide are forced to extend the use or reuse of their N95 FFRs [4]. Few options can be adopted for N95 FFR reuse. The US Food and Drug Administration (FDA) Emergency Use Authorization (EUA) guidance states that vaporized hydrogen peroxide gas plasma or hydrogen peroxide vapour (HPV) can be used for FFR decontamination if various conditions are satisfied, such as being free of visible damage and visual soil/contamination [5]. Due to post-processing form loss, vaporized hydrogen peroxide gas sterilization is not authorized for use with respirators containing cellulose-based or paper materials. Moreover, these techniques are not readily available in some middle- and low-income countries. Among the alternative methods, ultraviolet light germicidal irradiation (UVGI) is promising and is recommended by the US Centers for Disease Control and Prevention (CDC) [6].

UVGI-based viral deactivation is an easy-to-use solution, and its efficacy on N95 respirators has been confirmed by several studies [7, 8]. For example, some groups have previously investigated the use of the UVC emission available in biosafety cabinets for these tasks [9, 10]. Some FFR manufacturers have also provided their decontamination guidelines [11]. According to recent reviews, a minimum radiant exposure of 1 J·cm^-2^ is required for a consistent 3 log reduction in H1N1 or MS2 viral load [12]. However, a higher UV dose might be needed to achieve ≥3 log reduction of a non-enveloped virus, such as the more resistant adenoviruses, to obtain the EUA from the FDA as per their guidance [13]. FFR surface sterilization of more than 6 log inactivation for SARS-CoV-1 was reported for radiant exposures ranging from 600 mJ cm^-2^ to 3614.4 mJ cm^-2^ depending on the viral strain used [14, 15]. UVGI decontamination does not appear to influence the performance of the respirator, even after repeated UVC exposure [16, 17]. Lindsley et al. [17] showed for 1860 and 9210 3M respirators no change in flow resistance or filter burst strength at radiant exposures up to 120 J·cm^-2^, representing perhaps over 100 simulated disinfection cycles. Strap integrity remains an issue at higher radiant exposures, however, in practice, it is unlikely that N95 masks would be disinfected more than five times due to commonly-observed loss of fit at that point, given simple wear, independent of the disinfection process.

While filter efficacy of N95 FFRs has been studied [19], there are several knowledge gaps regarding N95 respirator decontamination using UVGI. A knowledge gap pertains to the understanding of light penetration through the fabric layers within the respirators. Previous studies did not provide a sub-layer resolution of the diffuse fluence, *H*, [mJ cm^-2^] within the respirator. We consider this to be important as it can help estimate the inactivation of the virus within the respirator. While processed FFRs were shown to have similar trapping abilities as naïve FFRs, even the reaerosolization of sub 1% of trapped viruses towards the room or the wearer are not acceptable [198]. This problem is amplified over repeat decontamination cycles. Another gap is the effect of the UV wavelength on the decontamination performance, as most earlier studies have focused on only 254 nm, which is emitted by traditional mercury arc-discharge lamps. Newer UV-LED systems can emit UV from 265 nm to 300 nm.

Through these studies, we aim to show respirator models suitable or unsuitable for further UVC decontamination. First, we determine the diffuse reflectance and transmittance for the FFRS fabric layers, derive the optical properties from 250nm to 350 nm and demonstrate the impact of the optical absorption and light scattering coefficients for the three fabric layers of seven common FFRs. The volume resolved fluence-rate [mW cm^-2^] distribution was studied using Monte Carlo simulations for two illumination schemes across the 254 to 280 nm range for a standard simplified geometry and a standardize irradiance [mW cm^-2^].

## 2 |EXPERIMENTAL

### 2.1 | FFR samples and preparation for optical measurements

The seven FFRs included 3M models 1805, 9105s (3M™ VFlex™), 1860, 8110s, 8210; 1870+, and 9210 (3M™ Aura™) (3M, St. Paul, MN, USA). They were obtained in April and May 2020 through the University Health Network and the University of Toronto. All respirators are cup or dome-shaped models and are approximated by a hemispherical shape for Monte Carlos simulations, independent of a permanent cup or a fold-flat design of the FFR.

The 3M 1805 FFR contains cellulose and is therefore not suitable for hydrogen peroxide-based decontamination. The 3M 1860 and 1870 FFRs do not contain cellulose knowingly; however, they are not certified cellulose-free. Nevertheless, the 1860 model is approved for HPV decontamination. The FFR design commonly comprises polyester-based materials for the outer and the inner shell, whereas the filter material is polypropylene or cellulose.

For the determination of the optical properties, 50 mm diameter samples were cut from the respirator and separated into three layers; for the outside layer, one sample was collected without printing and one with the test on the front. Figure 1 shows the prepared samples.

**FIGURE 1.**
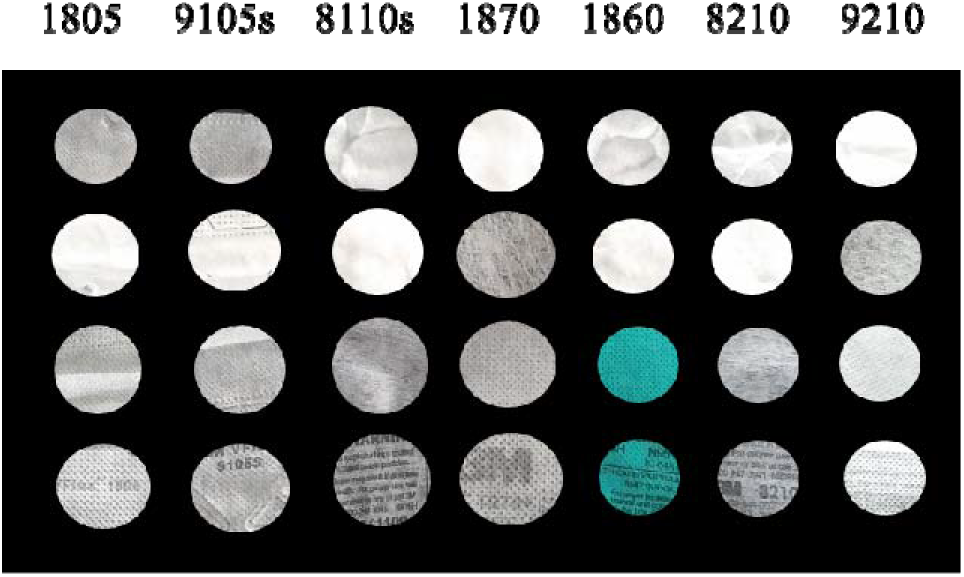
Respirator layer material samples of different FFRs (Model number indicated on top) for the determination of their optical properties. 1^st^ row Inside layer, 2^nd^ row Filter material, 3rd row outside layer without print and 4th row outside layer with print.

### 2.2 | Determination of FFR material optical properties

Optical properties were determined based on the approach of Scott Prahls [20], by quantifying the diffuse transmission and diffuse reflections using a single 6-inch integrating sphere (Labsphere, North Sutton NH, USA) with 1.25 inch (31.75 mm) entry and sample ports. The sphere’s surface reflectivity at 250nm and above was determined by exposing the side of the sphere with an open sample port and closed sample port as previously described [20, 21]. The measured sphere reflectivity is shown in Supplementary Figure S1. Data below 250 nm is unreliable and were excluded from further analysis. The resulting spectra suggest absorption bands within the Spectralon material or contaminations that may have accumulated over time.

The detector port was fitted with an optical fibre detector equipped with a cosine corrector (CC-3-UV, Ocean Optics, Rochester, NY, USA) and connected to a UV/VIS spectrophotometer (USB4F04880, Ocean Optics, Rochester, NY, USA) providing a spectral resolution of ~0.5 nm. Integration time was set to 6 secs, and 8 spectra were integrated with a boxcar smoothing of 2. While the long integration time resulted in PDA saturation at λ> 400 nm, no blooming was evident at the shorter wavelength of interest. Spectral collection and readout were performed with an Ocean Optics SpectraSuite software.

### 2.3 | Monte Carlo based light propagation in FFR

A medium pressure mercury lamp with collimator was used as the light source (Model: PS1-1-120, Calgon Carbon Corp., Moon Township, PA). The light beam was apertured to 17 mm before the integrating sphere’s entry port. Figure 2 shows an image of the physics setup. Background corrected transmission and reflection spectra were converted into diffuse transmission and reflection spectra. To apply the Inverse Adding-Doubling algorithm to extract the wavelength resolved absorption and reduced scattering coefficients [mm^-1^], the integrating sphere’s surface reflectivity needed to be determined [21]. The reflectivity was determined through off- axis measurements as proposed [22]. The outer filter and inner respirator layer thicknesses cannot be accurately measured due to different fabric compressibility, and the manufacturer did not provide fabric thickness details. Hence, thicknesses of 0.98 mm, 1.77 mm and 1.05 mm were assumed for the three layers and as fluence-rate-volume-histograms were determined for % volume rather than absolute volume and hence the assumption of the layer thickness.

**FIGURE 2.**
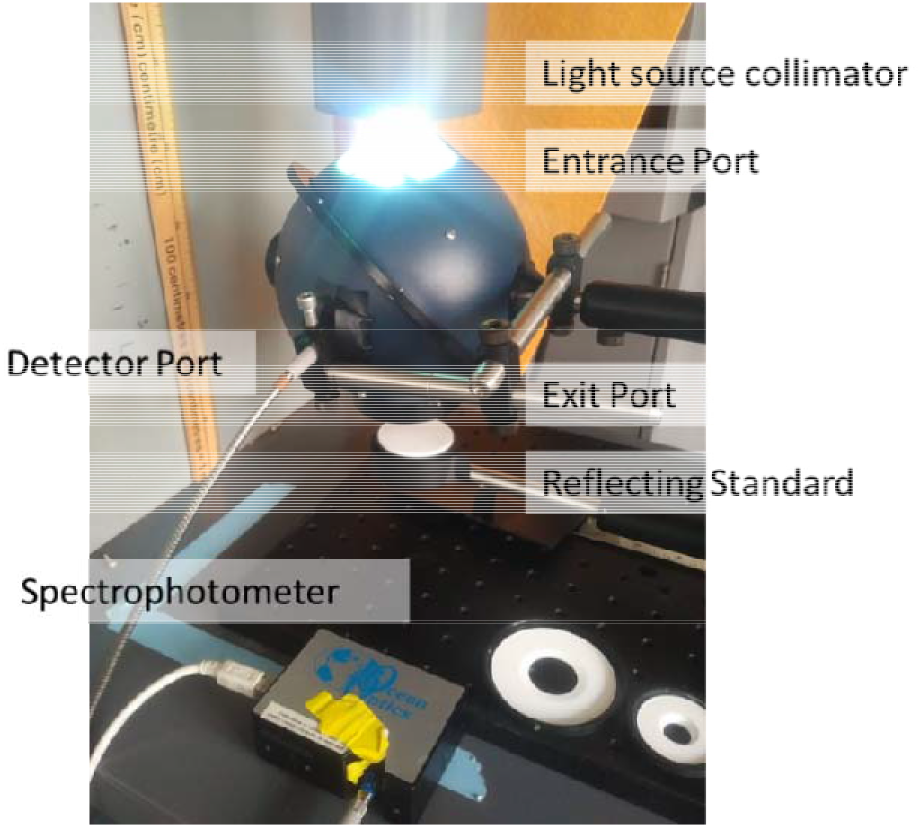
Physical image of the experimental setup.

Both the Inverse Adding-Doubling algorithm and the Monte Carlo simulations require the assignment of an average refractive index for each fabric layer [23], comprised of the textile fibres and air. The refractive index of polyester, a birefringent material, is given as 1.71 to 1.73 along the fibres and 1.53 to 1.54 perpendicular to them. The fabric density of Dacron® is 1.38 g cm^-3^. The initial photon incidence is perpendicular to the fibre axis. The mean flow pore diameters are estimated at 4-5 μm resulting in a packing density of 70% air volume. Hence, an average refractive index of 1.3 was assigned.

Polypropylene isotactic has a density of 0.91 g cm^-3^ and a refractive index of 1.492 with a fibre diameter of 1.2 μm. The mean flow pore diameter for non-thermal bonded fabrics is 1500 microns but is reduced to 10-15 microns by thermal calendaring [24]. Again, the overall fabric density is low, and an average refractive index of 1.15 was assigned.

To predict the minimum UVC fluence rate, Φ, at any point in an FFR, Monte Carlo based *in silico* photon propagation simulations were performed. Simulations were based on a standard 3D phantom fitted with the optical fabric properties derived above. Figure 3 shows an image of the hemispherical 3D phantom with an outer radius of 12 cm, comprised of the three layers, with 0.98, 1.77 and 1.05 mm representing the outer, filter and inner layer, respectively. The three-layer volumes are 24.84 cm^3^, 42.55 cm^3^ and 24.34 cm^3^, each divided into 129550, 118565 and 99801 elements. The average tetrahedral size per layer was 0.192 mm^3^, 0.358 mm^3^ and 0.244 mm^3^, accordingly. The hemispherical shape was chosen to include oblique UV radiation incidence on the FFR’s surface.

Three 1, 3, and 5 mm broad light-absorbing stripes were added to investigate the effect of the black printing on the fluence-rate inside the respirator. Additionally, a 5 mm wide and 2 mm thick object, simulating an aluminium nose piece, was added to the side of the hemisphere. In the 1^st^ set of simulations (A), illumination is via the outer shell with the respirators sitting supinely on an aluminium reflector and in the 2^nd^ set (B), illumination is via the inner layer with the FFR prone on the aluminium reflector. The absorption and reduced scattering coefficient of the aluminium pieces were 1 mm^-1^ and 20 mm^-1^, respectively, allowing for a high backscattering albedo.

Two sources were modelled. A large flat circular source of 12 cm diameter and centred 10 cm over the respirator’s apex when positioned supine and emitting normal to its surface simulated a very distant source. The second source represents a small 1 cm diameter photon emitting surface with a numerical aperture (NA) of 0.4 or 0.54, situated 10 cm over the supine or prone positioned respirator, respectively., The light sources were modelled as objects with photons emitted randomly over their surface triangles, either normal to the surface or within a user-defined solid angle to limit the FullMonte simulations resources [25].

10^8^ photon packets were launched in the FullMonte simulations [26], requiring less than 2 minutes, on an Intel(R) Xeon(R) CPU E5-2650 v4 running at 2.20 GHz equipped with 128 GB RAM when executing two threads on each of the 12 CPU cores. All sources were assumed to emit an irradiance of 1 mW cm^-2^ at their surface. The emitted photon packets represent the power emitted per source.

For the generation of the fluence-rate-volume-histograms (FVH), the photon weight absorbed by each volume element is corrected of its particular volume and converted to a local Φ (r), followed by sorting the tetrahedral in descending order of fluence-rate and plotted versus volume [%].

**FIGURE 3.**
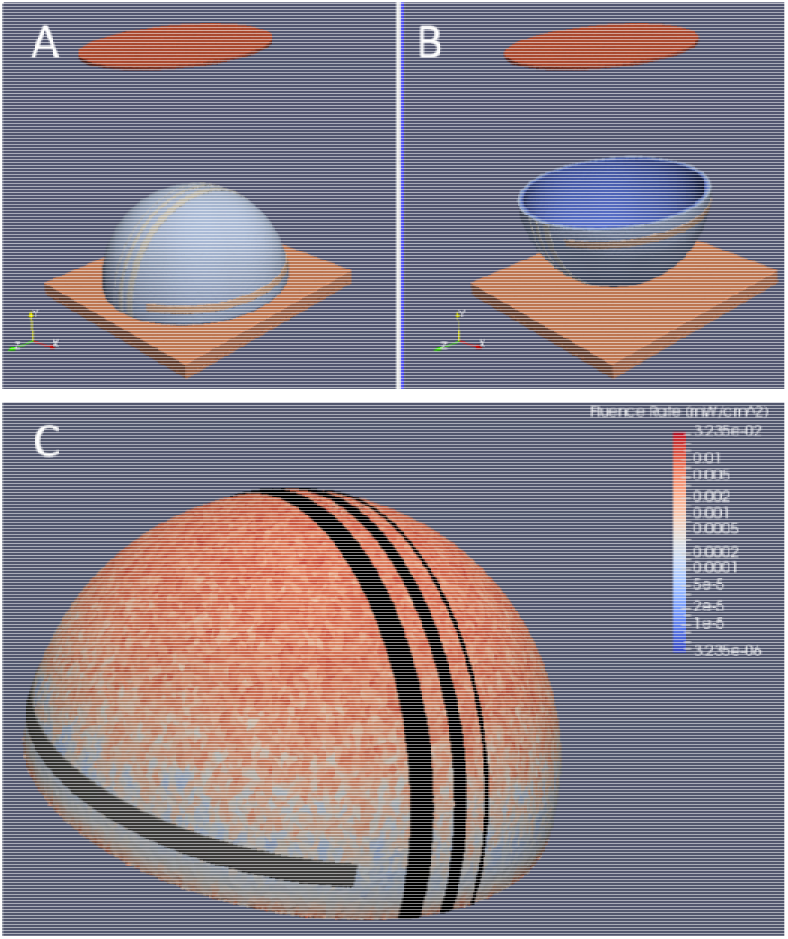
In silico phantom modelling setup used to simulate light propagation within the N95 FFRs. A) Illumination with a 12 cm diameter (red disc) via the outer layer and the hemisphere shaped respirator (blue) supine resting on a squared aluminium background. Light orange indicates the three different-sized absorbing layers and the horizontal aluminium strip. B) Illumination via the inner layer and the respirator prone on the aluminium layer. C) The surface irradiance is colour-coded for one respirator Model (3M 1805) at *λ* = 254 nm and a source emission of 1 mW cm^-2^

## 3 | RESULTS AND DISCUSSION

While various recent studies reported on surface irradiation variations on N95 FFR [27] or transmission of select wavelengths through the different respirator layer material [6], the reports were only for perpendicular illumination. These models considered only light absorption properties of the filtering fabrics rather than absorption and light scattering properties to achieve complete 3D UVC light, propagation models. The latter is required to determine the minimum attainable min within a respirator which was not previously reported, principally due to the unknown optical properties of the polyester, polypropylene, and cellulose fabrics utilized in the manufacturing processes and lack of suitable simulation platforms for light propagation emitting from large extended sources to the highly light-scattering materials.

While the fabrics comprise micron-sized fibres with varying degrees of packing densities, light propagation simulations such as FullMonte most often assume bulk optical properties. Here, the program also assumes isotropic optical properties and does not consider birefringent features present in polyester fibres. Figure S2 shows an example of the spectral conversions from raw spectra to fabric layer-specific diffuse transmittance and reflectance for the 3M 1860 and 9105s respirators.

Certain assumptions were made in determining the fabrics’ optical properties for the Monte Carlo simulation. When determining the integrating sphere’s reflectivity, the unknown reflection standard was iteratively changed until it was very close to the derived sphere reflectivity. Modulations of the sphere reflectivity due to potential absorption bands were not considered. The refractive index assigned to the outer shell and inner lining, as well as the filter materials, are estimates based on the fibre material and its packing density. The resulting layer-specific transmission values for the 1860 and 1870 respirators compare well to those determined using an actinometer-based technique (Dr Benoit Barbeau, Ecole Polytech Montreal, QB, Canada) and for data reported by Fisher et al. for 1860, 1870, and 8210 respirators. The absorption properties of the polyester and polypropylene have been determined previously. Polyester has a flat absorption in the UVA to the UVC region. However, there is a significant increase in absorbance below 245 nm. Polypropylene has considerable absorbance below 400 nm, albeit with little spectral shape. No sharp spectral absorption changes were seen in the 250 to 300 nm spectral range in our experiments. However, different materials have very different absorption coefficients as anticipated between polyester (low μ_a_) and polypropylene (high μ_a_).

Fabric fibres have a very high aspect ratio (>> 3.5), and their diameter is large compared to the wavelength of interest, and hence there aren’t large variations in the scattering coefficient. The scattering coefficient varies little once the form factor, ρ, is > 10. [28, 29] Based on a textile fibre diameter of 1.2 μm, ρ falls from 16.47 to 14.88 between 254 nm and 290 nm, respectively. This restricts the range for the scattering coefficient to within ~10%. Overall, the limited variations in the optical properties across the 46 nm range are not surprising.

Figure 4 presents the resulting reduced light scattering and absorption coefficients for the wavelengths of interest (254, 260, 280, and 290 nm). Table S1 lists all derived fabric optical properties for the three fabric layers at 254, 265, 280 and 290 nm in all 7 FFRs.

**FIGURE 4.**
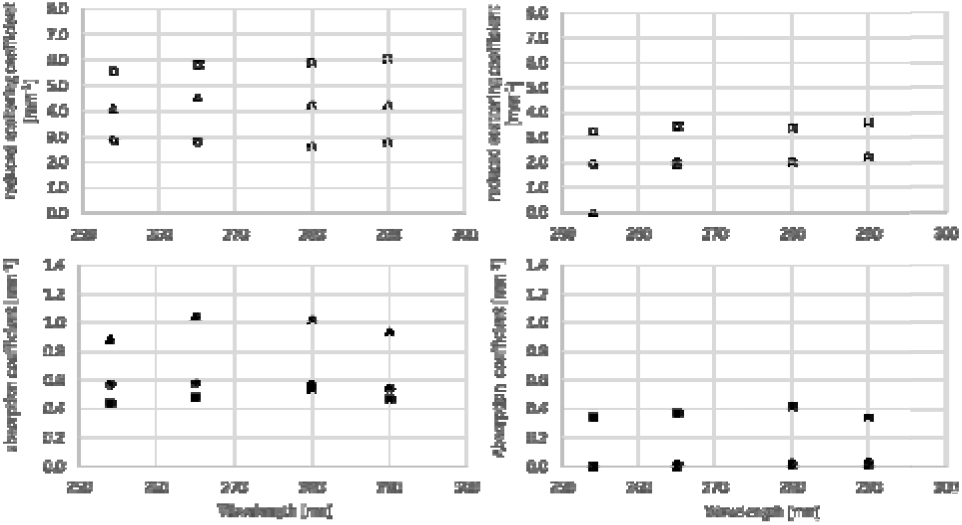
Wavelength-dependent reduced light scattering (top) and absorption (bottom) coefficients, for 3M 1870+ (left column) and 3M 9105s (right column) FFRs. Circles, triangles and squares stand for the outer, inner and filter layers, respectively.

Fluence-rate-volume-histograms were plotted for the dual photon sources, large collimated (Figure 5) and small divergent (Figure 6) emitting 254 nm, and the two illumination directions via the outer and the inner shells respectively, and for the combined illumination of an FFR 3M 1860 and the 3M 9105s FFRs at all four wavelengths. The FVHs for all 7 FFRs at the four simulated wavelengths are available online; see Data Availability Statement at the end of the manuscript. The FVH in Figures 5 and 6, two top rows, show that single side irradiation will not achieve complete volumetric decontamination. For illumination from both sides the oblique incidence of the UV light at the base of the FFR is the primary cause of low fluence rates in the material, see figure 3.

**FIGURE 5.**
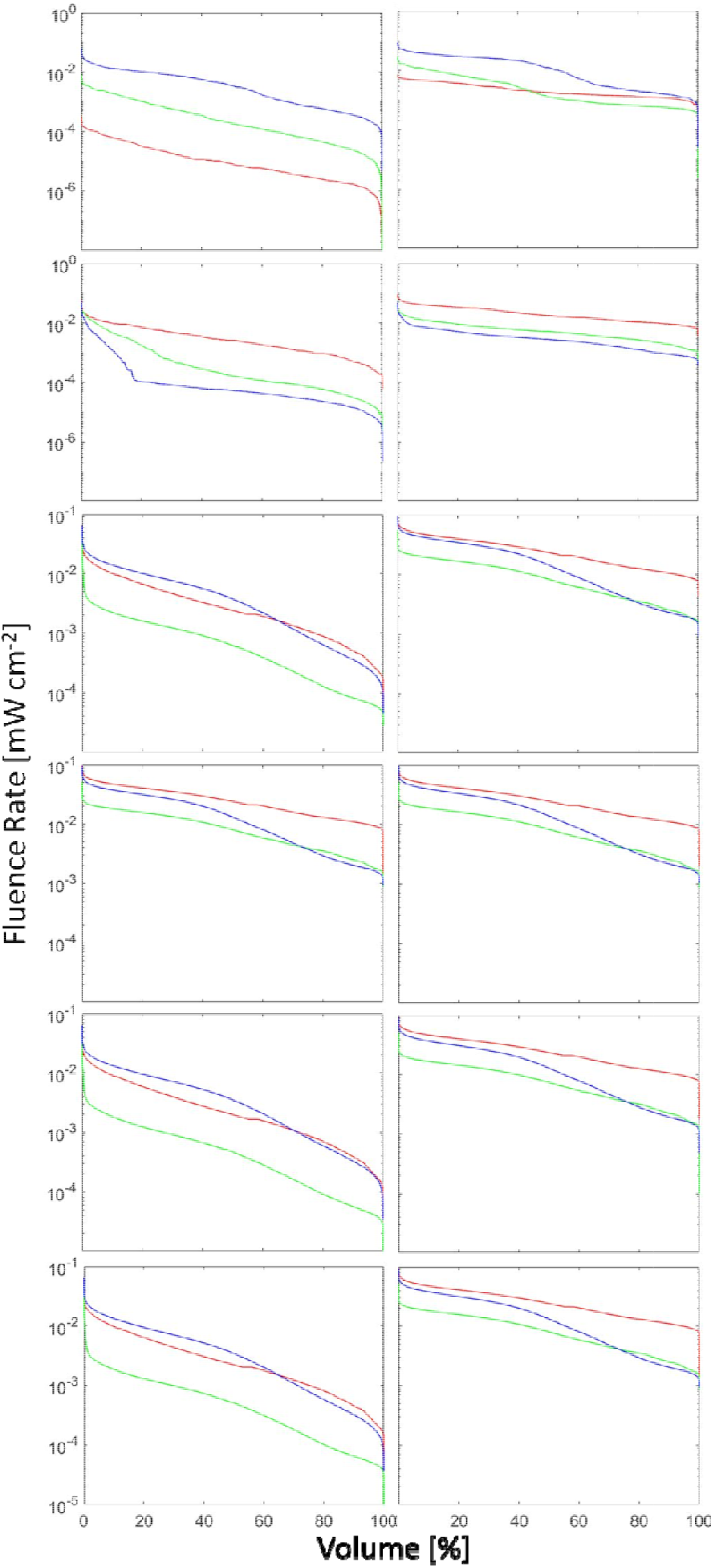
FVH for clean and dry FFRs: 1860 (left) and 9105s (right) for a 12 cm diameter, emitting 113.1 mW optical power. Outer shell blue, filter green and inner lining red. Top row λ.= 254 nm at (A) top irradiation, 2nd row λ = 254 nm at (B) bottom irradiation. Additive for (A) and (B) irradiation combined 3rd row λ = 254 nm 4th row λ = 265 nm, 5th row λ = 280 nm, 6th row λ = 290 nm.

**FIGURE 6.**
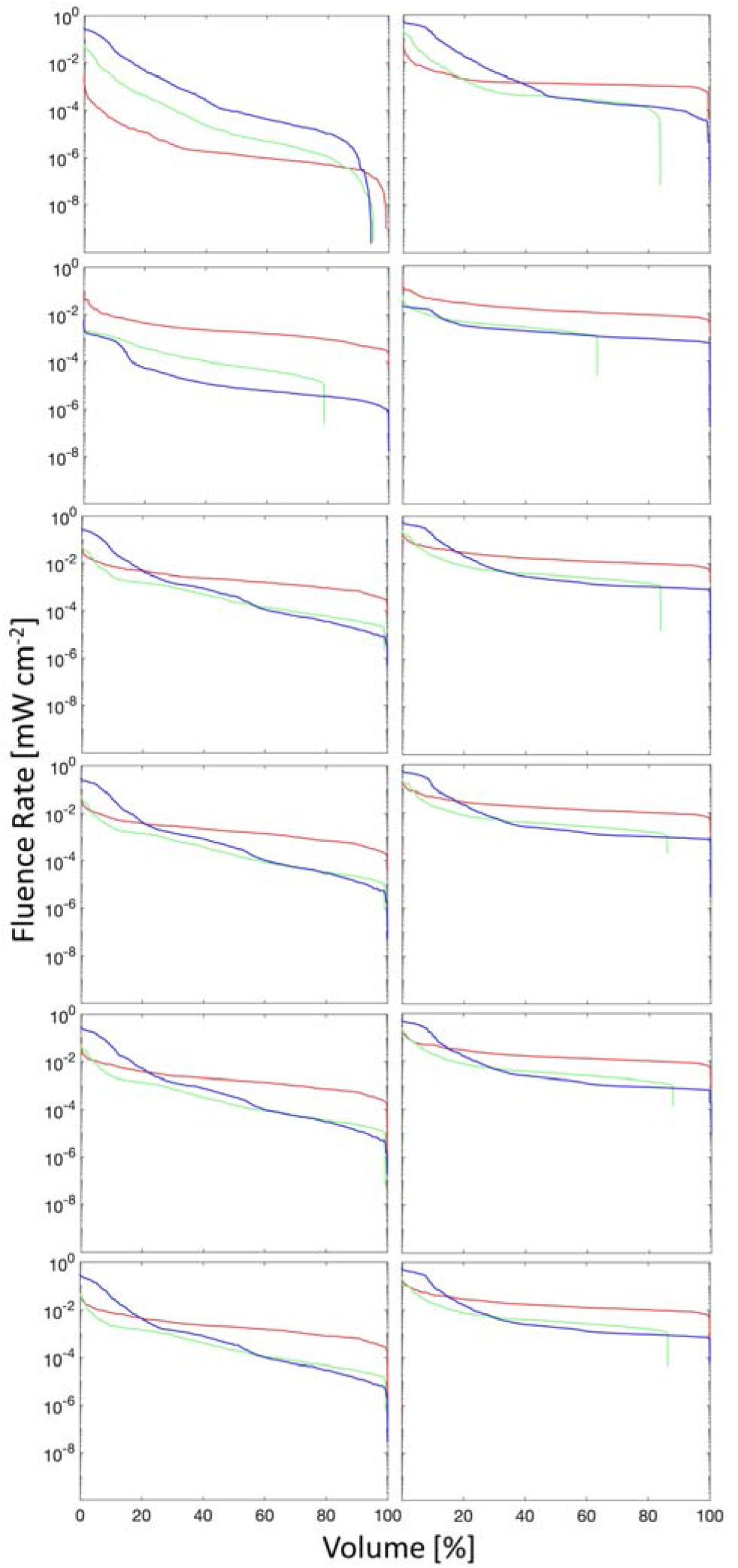
FVH for clean and dry FFRs: 1860 (left) and 9105s (right) for a 1 cm diameter emitting 0.78 mW optical power. Outer shell blue, filter green and inner lining red. Top row *λ*= 254 nm at (A) top irradiation, 2nd row = 254 nm at (B) bottom irradiation. Additive for (A) and (B) irradiation combined 3rd row = 254 nm 4th row *λ*= 265 nm, 5th row *λ*= 280 nm, 6th row *λ*= 290 nm. The total power was adjusted to be equal to the large source in Figure 5.

Not all tetrahedral received any photon-weight after launching 10^8^ photon-packets. Hence, the minimum required irradiance for 100% of the FFR volume was 0 for both the large and small source models. For the 3M 1860 FFR and 254 nm, increasing the number of photon-packets per simulation from 10^8^ to 10^10^ did not reduce the fraction of tetrahedrals with non-zero values significantly.See Table S2 for resulting calculation times and the number of tetrahedrals with 0 mW cm^-2^ fluence-rate while increasing the computational cost by close to 2 orders of magnitude. The resulting FVH are fundamentally identically, suggesting that a fluence-rates convergence within the dynamic range is present. The zero fluence rate tetrahedrals have small volumes and are found predominantely at the FFR edges opposite to the illumination source, see figure S2. Repeat simulations, n=3, for 1805 and 9105s FFRs, resulted in standard errors < 1 % for Φ_min_ > 0.25μcm^-2^.

For a nominal source emission of 1 mW cm^-1^, the lowest fluence rate, €>, at 99% or 95% of the FFR volume following the combined illumination via outer and inner shell for the four simulated wavelengths, are summarized in Tables 1 and 2, respectively.

**TABLE 1.** Minimal attainable fluence rate O_min_ for 99 % of the FFR volume for two modelled light illumination geometries, each emitting 1 mW cm^-2^ for 4 different wavelengths from 254 nm to 290 nm.

| FFR | Large source<br>$\Phi_{\min} [\mu\text{W cm}^{-2}]$ | | | | Small source<br>$\Phi_{\min} [\text{nW cm}^{-2}]$ | | | |
| --- | --- | --- | --- | --- | --- | --- | --- | --- |
|  | 254 | 265 | 280 | 290 | 254 | 265 | 280 | 290 |
| 1805 | 0.236 | 0.218 | 0.193 | 0.227 | 0.164 | 0.142 | 0.122 | 0.154 |
| 1860 | 0.053 | 0.036 | 0.035 | 0.041 | 0.002 | 0.001 | 0.001 | 0.001 |
| 1870+ | 0.286 | 0.234 | 0.273 | 0.341 | 0.063 | 0.051 | 0.062 | 0.083 |
| 8110s | 0.034 | 0.058 | 0.071 | 0.041 | 0.001 | 0.004 | 0.008 | 0.001 |
| 8210 | 0.073 | 0.047 | 0.045 | 0.061 | 0.005 | 0.001 | 0.001 | 0.003 |
| 9105s | 1.704 | 1.588 | 1.38 | 1.47 | 1.660 | 1.449 | 1.121 | 1.266 |
| 9210 | 0.346 | 0.293 | 0.297 | 0.358 | 0.120 | 0.092 | 0.096 | 0.129 |

**TABLE 2.** Minimal attainable fluence rate Φ_min_ for 95 % of the FFR volume for two modelled light illumination geometries, each emitting 1 mW cm^-2^ for 4 different wavelengths from 254 nm to 290 nm.

| FFR | Larg source<br>$\Phi_{\min} [\mu\text{W cm}^{-2}]$ | | | | Small source<br>$\Phi_{\min} [\text{nW cm}^{-2}]$ | | | |
| --- | --- | --- | --- | --- | --- | --- | --- | --- |
|  | 254 | 265 | 280 | 290 | 254 | 265 | 280 | 290 |
| 1805 | 0.339 | 0.320 | 0.291 | 0.330 | 0.266 | 0.242 | 0.215 | 0.251 |
| 1860 | 0.067 | 0.047 | 0.045 | 0.053 | 0.004 | 0.001 | 0.001 | 0.002 |
| 1870+ | 0.367 | 0.301 | 0.344 | 0.416 | 0.084 | 0.086 | 0.119 | 0.144 |
| 8110s | 0.049 | 0.083 | 0.098 | 0.057 | 0.001 | 0.008 | 0.013 | 0.002 |
| 8210 | 0.093 | 0.065 | 0.061 | 0.080 | 0.007 | 0.003 | 0.002 | 0.005 |
| 9105s | 2.016 | 1.896 | 1.680 | 1.775 | 2.071 | 1.849 | 1.478 | 1.607 |
| 9210 | 0.422 | 0.361 | 0.362 | 0.435 | 0.180 | 0.139 | 0.145 | 0.189 |

Manufactured printing on the outer shell layer does not influence the overall decontamination of the respirator due to the small added absorption relative to the fabric’s absorption in the ultraviolet region of the optical spectrum. However, personalization of FFRs with Sharpies and other penetrating ink-based markers needs to be avoided, as are cosmetic cremes and lipsticks. Considering a ten v/v% contamination of the outer layer with patient saliva or the inner layer with the bearer’s saliva would increase the absorption coefficient by 0.085 mm^-1^ (Prof. Benoit Barbeau, Polytechnique Montreal, personal communication), comparable to the printing on the front shell, and thus would not impact the light distribution significantly.

The aluminium nose bridge did not cause significant shadowing effects for the simulated illuminations as it was not perpendicular to the direction of incidence. However, when light sources are coming sideways, significant shadowing will occur, possibly preventing the achievement of FDA guideline decontamination. There are substantial differences in the light scattering and absorption properties between the fabrics.

Using a small light source is not desirable as excessively long treatment times are required to achieve the necessary fluence, *H*, over at least 99% of some FFRs’ volume for UVGI. This is independent of the 144 times lower total power emitted by the smaller source.

However, the higher irradiance alone does not explain the lower Φ in some volume elements of the hemisphere. The low Φ is due to a partial surface shielding of the FFR by its form and oblique incidence angles of the photons, see Figure 3. Conversely, the 99% data is reliable as sufficient absorption events were recorded. The influence of uncertainties in the optical property is limited. Increasing μ_s_ by 10% in all three layers did not affect the Φ_min_ shown in Tables 1 and 2 for the 3M 1805 FFR and reduced Φ_min_ between 4.5% and 5.3% for the 3M 9105s FFR.

The required irradiation times, t, required to achieve the minimum fluence,, for UVGI of SARS-CoV-2 or other targets are calculated by 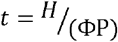, with P representing the irradiance in the mask plane. For example, to reach 65 mJ cm^-2^ for MS2 inactivation [30] by a large collimated source emitting 1 mW cm^2^ of 254 nm would require over 32 times longer for a 3M 1860 FFR than a 3M 9105s.

It is clear that using large-area flat illumination, without considering the FFR’s shape, will always result in very long exposure times to attain the minimum required fluence throughout the mask volume in a reasonable time. They can be significantly reduced by optimizing the shape and position of the light source for a given FFR shape, Duckbill versus parabolic cup shape, utilizing available inverse optimization algorithms [31]. Additionally, higher emission light sources are also required to bring the decontamination times into the one minute range to achieve acceptance of UVGI.

The suitability of the FFRs for UVGI depends primarily varies widely for 4 of the tested models. Suitable M3 models include 1805, 1870+, 9210 and 9105s, where the latter would require the shortest illumination times. Three of the tested FFRs, 1860, 8110s and 8210, are less suitable for UVGI. Achieving the fluence-rate needed for a 6 log viral or bacterial inactivation, as currently defined by the FDA, would result in an excessive irradiation time for the investigated conditions. Reduction of the irradiation times can be achieved, by optimizing the source placement in actual devices, also considering the reflectivity of irradiation chambers. However, the high photon absorption and scattering coefficient, of some of the employed fabrics remain a principal challenge.

To physically prove the potential of UVGI for a given FFR for a given illumination geometry modifications to the *in silico* modes are required, such as its actual physical shape and the total number of tetrahedrals must exceed 10^6^ to demonstrate a 6 log reduction of particular specified test organisms stated by the current (June 10^th^, 2020) Guidance for Industry and Food and Drug Administration Staff for EUA for PPE Decontamination [32]. For the physical implementation of UVGI, the fluence-rates would likely be one to two orders of magnitude higher since decontamination devices would be constructed to provide reasonably uniform coverage of UV across the entire outer mask surface. However, the difference in FFR suitability for UVGI is not affected by either of these shortcomings, and each FFR model needs to be independently evaluated. Comparisons with actual biological experiments evaluating the complete volumetric decontamination of some of these FFRs are highly desirable.

## 4 | CONCLUSION

UVGI is a reasonable approach for FFR decontamination to extend a respirator’s usable lifetime when supply chains are restricted during public health emergencies. Both the investment costs and environmental impact are low. Operator exposure to harmful UV radiation and ozone can be minimized by simple technical safety measures. However, to achieve consistent, fast and complete decontamination, the photon-source positions need to be paired to the respirator shape, and the optical properties of the FFR model to be irradiated needs to be determined at the UVGI wavelength, to establish the minimum required exposure time based on photon distribution simulations as described here.

## Data Availability

The raw data and all fluence-rate-volume-histogram https://www.eecg.utoronto.ca/~vaughn/fullmonte/n95_decon/n95_decon.html

## ACKNOWLEDGMENTS

Funding for this study was provided through the Ontario Ministry of Economic Development and Trade, through the Ontario Research Fund Grant # ORF RE08-022 and Princess Margaret Cancer Centre, the Princess Margaret Cancer Foundation, and Ontario Ministry of Health, NSERC/Intel Industrial Research Chair in Programmable Silicon IRC:428842-16 and an IBM Faculty Research Award. The authors thank Theralase Technology Inc., Toronto for the loan of the integrating sphere.

## AUTHOR CONTRIBUTIONS

L.L. designed the overall experiments and collected primary data supported by A. N., A.M. and M.W. executed the Monte Carlo Simulations and generated the FVHs, F.S. and T. Y-S, modified FullMonte, so it was suitable for the task, A. S. A., C.W, F.X.G, and R.H provided the UV decontamination context. Everybody contributed to the completion of the manuscript.

## DATA AVAILABILITY STATEMENT

**ABBREVIATIONS**
FFR: Filtered Face Respirators
FDA: US Food and Drug Administration,
EUA: Emergency Use Authorization, Hydrogen peroxide vapour
HPV: ultraviolet light germicidal irradiation
UVGI: US Centers for Disease Control and Prevention
CDC: fluence rate-volume-histograms FVH,

## SUPPORTING INFORMATION

Figure S1 shows the diffuse reflectivity of the integrating sphere in the 250 to 350 nm range. Figure 2 shows the location of zero value tetraheadral for a 3M 1860 FFRs. Table S1 lists the determined fabric optical properties, and μ_s_’, for three fabric layers of the seven evaluated FFRs at 254, 265, 280 and 290 nm.

## AUTHOR BIOGRAPHIES

**Prof. Dr. Lothar Lilge** received a Diploma in Exp. Physics from the Johann Wolfgang Goethe University, Frankfurt and a PhD from the Westfaehlische Wilhelms University in Muenster Germany. Further training was at the Wellman Laboratories in Photomedicine, Boston, USA and McMaster University, Hamilton, Canada. He is Senior Scientist at University Health Network and Faculty in the Department of Medical Biophysics at UofT. He currently works on various aspects of Photodynamic Therapy, and optical breast cancer risk assessment.

**Angelica Manalac** received her BSc degree in Medical Physics from McMaster University, Hamilton, Canada in 2020. She is an incoming Master’s student in the Medical Biophysics graduate program at the University of Toronto, Toronto, Canada.

**Madrigal Weersink** is an undergraduate student in the Faculty of Engineering at McMaster University, Hamilton, Canada.

**Dr. Fynn Schwiegelshohn** received his MS degree from the Department of Electrical and Information Technology, Technical University of Dortmund, Germany in 2007. He received his Ph.D. in reconfigurable computing from the Ruhr-University Bochum, Germany. His research focuses on exploring algorithm accelerations for biomedical research and applications.

**Tanner Young-Schultz** received his BASc degree from the Department of Electrical Computer Engineering at the University of Toronto. He is currently working towards his MASc at the University of Toronto focusing on exploring algorithm acceleration using CPUs, GPUs and FPGAs.

**Dr. Abdallatif Satti Abdalrhman** is a Postdoctoral Fellow in the Department of Civil and Mineral Engineering at the University of Toronto. He received his B.Sc. degree in Chemical Engineering from the University of Khartoum, M.Sc. in Environmental Engineering from Tsinghua University, and PhD in Environmental Engineering from the University of Alberta.

**Dr. Chengjin Wang** received his Bachelor’s and Master’s degrees from Tongji University and his PhD degree in Environmental Engineering from the University of Alberta. He is working as a Postdoctoral Fellow at the University of Toronto on advanced oxidation processes in water treatment.

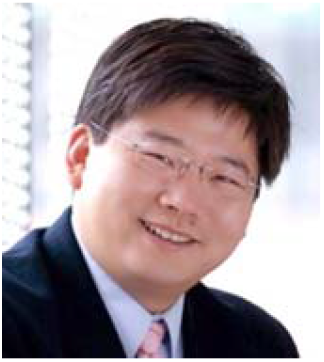

**Aldrich Ngan** received his Bachelor’s and Master’s degrees in Chemical Engineering from the University of Toronto. He is currently a Ph.D. student at the University of Toronto studying the removal of inorganic contaminants in industrial wastewaters.

**Prof. Frank Gu** is a Professor in the Department of Chemical Engineering and Applied Chemistry at the University of Toronto. He holds an NSERC Senior Industrial Research Chair in Nanotechnology Engineering. He completed his PhD in chemical engineering at Queen’s University in Kingston, Canada. His research group focuses on combining chemical engineering principles and nanotechnology to tailor the material design for life sciences and environmental remediation applications.

**Prof. Dr. Vaughn Betz** is a Professor and the NSERC/Intel Industrial Research Chair in Programmable Silicon in the Electrical and Computer Engineering Department at the University of Toronto. His research focii include optimization algorithms, biophotonic simulation, and the creation of hardware accelerators to speed up simulations.

**Prof. Dr. Ron Hofmann** is an NSERC Associate Industrial Research Chair in Advanced Drinking Water Technologies in the Department of Civil and Mineral Engineering at the University of Toronto, with a focus on UV disinfection.

